# Can dental EHR data support a learning health system? Completeness, consistency, and accuracy of structured versus narrative diagnoses

**DOI:** 10.64898/2026.11.04.26367297

**Authors:** Yao-Shun Chuang, Guo-Hao Lin, Chun-Teh Lee, Ryan Brandon, Muhammad F. Walji, Xiaoqian Jiang, Oluwabunmi Tokede

**Author notes:** **Corresponding Author**: Oluwabunmi Tokede, DDS, MPH, DMSc, Department of Diagnostic and Biomedical Sciences, University of Texas at Houston, Health Science Center, 7500 Cambridge Street Houston, TX 77054. **Author Contributions:** Assign CRediT roles per author, e.g.: Y.-S.C.: Conceptualization, Methodology, Software, Writing – original draft; G.-H.L. / C.-T.L.: Investigation, Validation (gold-standard review); R.B.: Data curation; M.F.W. / X.J.: Supervision, Writing – review C editing; O.T.: Conceptualization, Supervision, Funding acquisition, Writing – review C editing. **Ethics Approval and Consent:** Institutional Review Board approval was obtained. Given the retrospective design using pre-existing data, a waiver of patient informed consent was granted.

## Abstract

**Introduction:** Electronic health record (EHR) data increasingly drive quality monitoring and research, yet their reliability in dentistry is uncertain. We compared the completeness, consistency, and accuracy of periodontal diagnoses captured in two different formats (structured fields versus narrative notes) across two academic dental centers sharing the same EHR platform and diagnostic terminology.

**Methods:** In this retrospective study of qualifying periodontal encounters (2021), structured diagnoses were extracted by SQL and narrative diagnoses by a fine-tuned RoBERTa NLP model. A random sample (∼20%) was independently reviewed by two board-certified periodontists, with disagreements resolved by consensus, to form the gold standard. Outcomes were completeness (presence of a diagnosis), consistency (agreement between formats), and accuracy (agreement with the gold standard), with ordinal periodontal stage compared using quadratic weighted κ.

**Results:** Across 4,542 encounters, completeness varied widely by site and format (34.7%–84.5%), and neither format was consistently more complete; at one site, over one-third of encounters (35.8%) had no documented diagnosis in either format. Among dual-documented encounters, agreement between formats was substantial but imperfect (κ = 0.86 and 0.80). Accuracy against the gold standard was modest for both sources (all near 50%), with narrative notes numerically higher at both sites and favored by the severity-weighted κ, though raw differences were not significant. Pooled across sites, the formats diverged in 14.8% of dual-documented encounters, 8.4% by two or more stages, with the structured field carrying the milder stage in 67.9% of disagreements.

**Conclusions:** Structured and unstructured periodontal diagnoses each matched expert adjudication only about half the time, and neither was consistently more complete. Narrative notes held a small severity-weighted accuracy advantage, and coded fields more often carried the milder stage when the two disagreed. These convergent, cross-site patterns appear systematic rather than site-specific and expose latent risks for analytics and decision support. Improving dental EHR data quality will require workflows that combine the computability of coded entries with the fidelity of free text.

## Introduction

Electronic health records (EHRs) have become a cornerstone of modern healthcare delivery, serving both as repositories of patient information and as tools to improve care coordination, quality of care, and patient outcomes. Documentation within EHRs typically falls along a spectrum, from structured (coded fields, templates, and standardized terminologies) to unstructured (narrative free text, dictated notes, and clinician comments). Understanding the strengths, limitations, and interplay of these two forms is critical for optimizing data usability, supporting clinical workflow, and enabling secondary uses such as research, quality measurement, and public health surveillance.

Structured documentation^1^ refers to data recorded in pre-defined formats such as drop-down menus, checkboxes, and templates, often using standardized terminologies such as SNOMED CT^2^, the Systematized Nomenclature for Dental Diagnostic System (SNODDS)^3^, ICD^4^, and CDT. Because these elements are coded and standardized, they reduce variability and can be readily queried, aggregated, and analyzed; they also facilitate clinical decision support (CDS) and interoperability across disparate health information systems. These same features, however, carry costs. Recording findings into rigid templates can be time-consuming and forces clinicians to adapt their workflow to the EHR design rather than their own clinical reasoning, increasing cognitive load and reducing face-to-face patient interaction^5,6^. Structured fields may also fail to capture the subtleties of a patient’s condition, with important clinical narratives, rationales for decisions, and unique circumstances lost or oversimplified^7^.

Unstructured documentation, by contrast, appears as narrative text produced through typing, speech recognition, or transcription^8^. It offers greater flexibility and expressiveness, often mirroring the way clinicians think and communicate, and supports the recording of subtle details, rationales, and complex decision-making that do not fit neatly into pre-defined categories; many clinicians therefore find free-text entry more natural and less disruptive to the patient encounter. These same narratives, however, are noisy and heterogeneous, and not readily machine-processable without natural language processing (NLP) or text-mining techniques^6,9^. Variation in language, abbreviations, misspellings, and documentation style limits the comparability and reliability of narrative data, and the absence of inherent structure makes it difficult to de-identify, share, integrate, and repurpose across systems, impeding large-scale analytics and decision support^10^.

Regardless of format, the secondary use of EHR data offers substantial potential to reduce research costs, enable patient-centered studies, and accelerate medical discovery^11^. This premise underlies the learning health system, in which data generated during routine care are translated into knowledge and returned to the point of care as improved practice^12^. Such a cycle is only as reliable as the documentation that seeds it: what is mis-recorded, or not recorded at all, at the chairside propagates into every inference the system subsequently draws. Yet this reuse faces persistent challenges, particularly around data quality and suitability for research: because clinical care and research have differing priorities, clinical data are often not recorded with the precision and rigor of data collected for research^13^, and prior assessments of EHR data quality have yielded highly variable results^14^. Among the dimensions proposed for evaluating suitability for secondary use, completeness, accuracy, and consistency are central^14^. Completeness refers to the extent to which the data required for a given purpose are present and adequately recorded, including the presence or absence of missing values^15,16^; accuracy is the degree to which recorded values truly represent the real-world events or conditions they describe^17,18^; and consistency is the degree of agreement among data recorded for the same patient across sources or formats.

Recent advances in artificial intelligence (AI) and natural language processing have markedly improved the ability to process and analyze unstructured clinical data, creating new opportunities to incorporate narrative information into research and, potentially, clinical use. Dentistry is actively building the infrastructure such a cycle requires, including standardized diagnostic terminologies, integrated diagnostic entry, and multi-institutional data networks^3,19^. Yet these documentation trade-offs and their consequences for data quality remain largely unexamined in dentistry, where structured diagnostic terminologies are still relatively new. In this study, we comparatively evaluate the quality of structured and unstructured data within a dental EHR and assess its fitness for the secondary uses on which a learning health system depends, focusing on the dimensions of completeness, accuracy, and consistency. We use periodontal diagnosis as a representative case to evaluate the reliability of dental EHR data for the analytics, quality measurement, and decision support through which such a system closes the loop.

## Research Interests

Using periodontal diagnosis as a representative case, and analyzing two academic dental centers in parallel to distinguish systematic patterns from site-specific ones, we asked whether structured (coded) and unstructured (narrative) documentation differ in three dimensions of data quality:

- Completeness: Among encounters for which a periodontal diagnosis was clinically indicated, how often is a diagnosis documented in structured fields versus narrative notes, and does either format capture diagnoses more completely?
- Consistency: Among encounters documented in both formats, how well do the structured and narrative diagnoses agree, and when they disagree, in what magnitude and direction?
- Accuracy: Among encounters with a documented diagnosis, how often does each format match an expert gold-standard diagnosis, and does either format align more closely with expert adjudication?

## Methods

### Ethical Considerations

Institutional Review Board approval was obtained. Given its retrospective design using pre-existing data, a waiver of patient informed consent was granted. All data were de-identified, stored on secured institutional servers, and accessible only to authorized study personnel.

### Study Design and Setting

This retrospective study was conducted at two academic dental institutions that share the same EHR platform (axiUm, Exan, Vancouver, Canada) and the same integrated diagnostic terminology, the Systematized Nomenclature for Dental Diagnostic System. The primary objective was to compare the completeness and accuracy of periodontal diagnoses recorded in structured EHR fields against those documented as unstructured free text in the progress notes. Where a diagnosis was recorded in both sources for the same patient at the same appointment, we additionally evaluated the consistency between the two formats. The two sites were analyzed in parallel rather than against each other: instead of ranking institutions or testing for inter-site differences, we asked whether the same structured-versus-unstructured patterns recurred across independent settings on a common platform, since convergent findings would indicate systematic features of the two documentation formats rather than the idiosyncrasies of one institution. Our overall approach is summarized in Figure 1.

**Figure 1:**
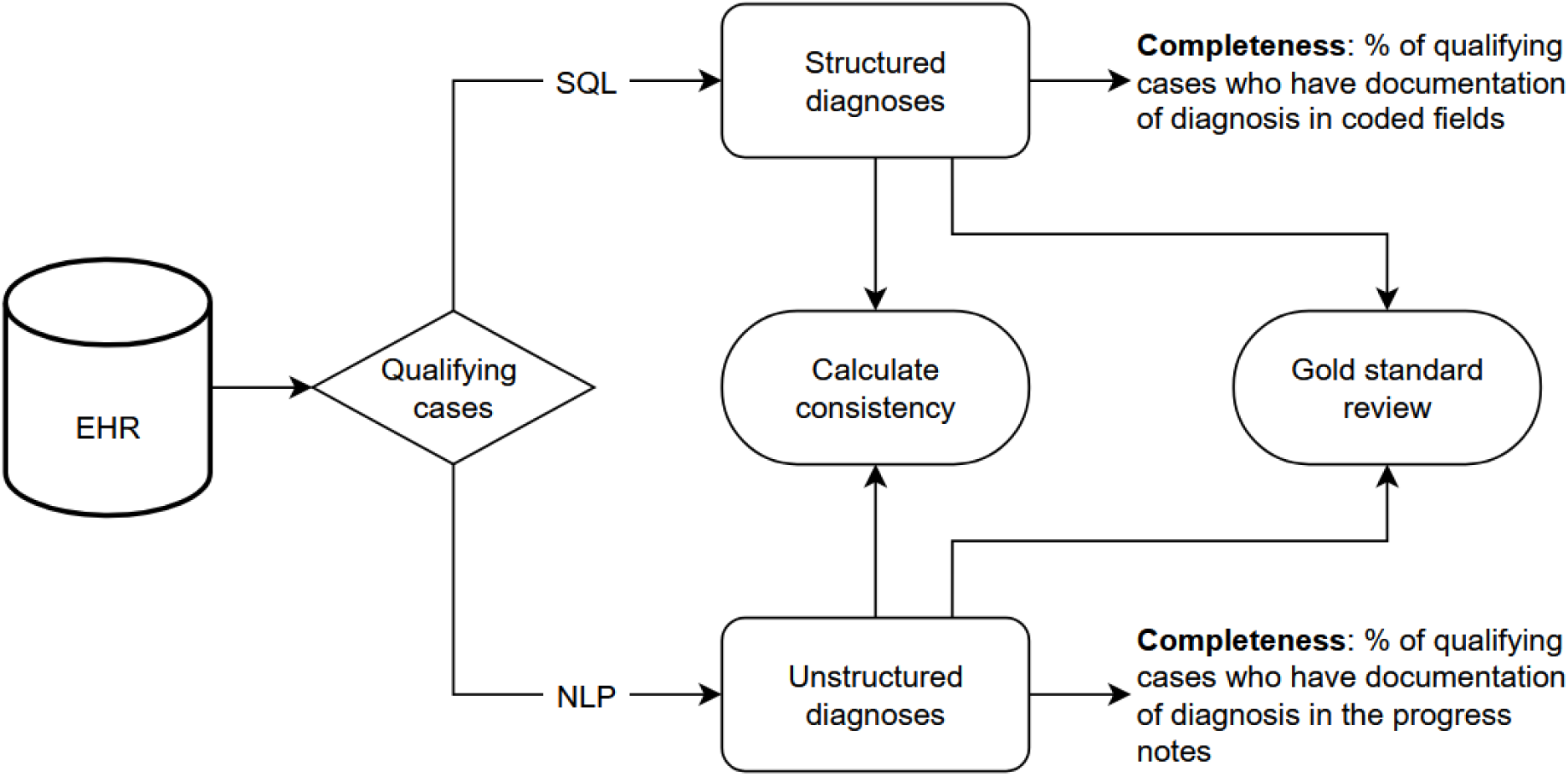
Study Overview.

### Data Source and Population

We identified a cohort of patients who received a qualifying periodontal examination during the study period (January 1, 2021, to December 31, 2021). Eligibility criteria included having at least one encounter in which a periodontal evaluation or treatment was documented. Cases were limited to adult patients with a minimum of 10 natural teeth present. For accuracy, cases additionally had to have comprehensive periodontal charting including pocket depth, clinical attachment level, and the distance from the free gingival margin to the cemento-enamel junction, and recent bitewing radiographs (within 6 months). These additional criteria defined the gold-standard–eligible subset used for the accuracy analysis, which was necessarily smaller than the full completeness cohort.

### Structured Data Extraction

Structured data pertaining to periodontal diagnoses were extracted using SQL scripts that queried specific EHR fields and/or coded diagnoses SNODDS -that uses the American Academy of Periodontology 2017 World Workshop periodontal disease diagnostic criteria^20^. Our multidisciplinary team of clinicians, informaticians, and data specialists reviewed the EHR data model to pinpoint fields capturing structured periodontal diagnosis information. Our team has extensive experience extracting data from EHRs and has iteratively developed custom SQL scripts. These scripts were validated through manual review by comparing query outputs with patient encounters in the EHR, as described in our previous work^19^.

### Unstructured Data Extraction and NLP Processing

For the same encounters, the free-text clinical notes associated with each visit were extracted to capture periodontal diagnoses documented in narrative form. Notes were preprocessed by removing extraneous formatting characters. To develop the extraction model, a representative subset of 45 de-identified notes was selected and manually labeled by the study team. Each labeled note then served as a template from which the OpenAI GPT-4 API generated 10 synthetic notes, yielding 450 synthetic notes for model training. Prompts were designed to preserve the clinical nuances specific to periodontal documentation, and sentence structure was varied to mirror real-world charting; all generation was performed on the HIPAA-compliant Azure OpenAI platform. These synthetic notes were used exclusively to fine-tune the RoBERTa^21^ (base) model and were not part of the analytic cohort. The fine-tuned model was then applied to the real clinical notes in the study cohort to identify and categorize periodontal diagnoses along the dimensions of stage, grade, and extent. Model performance was evaluated against manually labeled real notes, with full development and validation methods reported separately^22^. On the gold-standard evaluation sets from both sites, the fine-tuned model achieved weighted-average F1 scores of 0.84–0.99 across the five diagnostic dimensions (periodontal status, stage, grade, extent, and subtype), with the lowest performance on extent and the highest on stage, grade, and subtype.

### Gold-Standard (GS) Review

A reference set was generated from a random sample (about 20% of the sampling frame). Two primary reviewers independently reviewed the included charts and then discussed cases in which their assessments varied to achieve a consensus in every case. Both are clinician-scientists with an in-depth knowledge in the etiology and treatment of periodontal disease. They are certified by the American Board of Periodontology and have extensive experience (>15 years) in providing complex patient care and conducting clinical research as full-time faculty. The reviewer-assessed diagnoses thereby created a gold-standard dataset against which the structured and unstructured diagnoses could be compared.

### Outcome and Statistical Analysis

We evaluated three dimensions of data quality: completeness, consistency, and accuracy. Because periodontal diagnoses are ordinal (health/gingivitis through Stage IV), agreement was quantified throughout with quadratic weighted κ, which penalizes disagreements in proportion to the square of the number of stages separating them, so that larger stage discrepancies, which carry disproportionately greater clinical consequence, contribute more heavily to the coefficient than single-stage discrepancies.

a. Completeness was the proportion of qualifying encounters, those for which a periodontal diagnosis was clinically indicated, with a documented diagnosis in a given data source. It was computed separately for structured and unstructured data across the full qualifying cohort, and we cross-tabulated how often the two formats co-occurred (both present, structured only, unstructured only, or neither).
b. Consistency was the agreement in assigned periodontal stage among encounters documented in both formats, reported as raw agreement (the proportion of concordant encounters) and quadratic weighted κ. For discordant encounters, we summarized the magnitude (single-stage versus ≥2-stage mismatch) and direction (whether the structured field assigned a milder or more severe stage than the narrative note) of disagreement.
c. Accuracy was the proportion of documented diagnoses whose recorded stage matched the single expert-adjudicated diagnosis for that chart, computed separately for structured and unstructured data within the gold-standard–eligible subset, with 95% confidence intervals and quadratic weighted κ. Both formats were scored against the same fixed set of adjudicated charts, but a format could be scored only where it carried a diagnosis, so the two estimates drew on overlapping but not identical subsets. The formats were therefore compared as independent proportions, with the between-format difference in raw agreement reported alongside its 95% confidence interval rather than a paired test.

All analyses were performed in R (RStudio, Inc., Boston, MA).

## Results

A total of 4,542 encounters met the inclusion criteria, comprising 2,055 at Site 1 and 2,487 at Site 2 (Table 1). A structured periodontal diagnosis was recorded in 1,650 encounters (80.3%) at Site 1 and 1,253 (50.4%) at Site 2, while an unstructured diagnosis was present in 1,736 (84.5%) and 863 (34.7%), respectively. The more complete format differed by site: at Site 1, unstructured notes carried a diagnosis somewhat more often than structured fields (84.5% vs 80.3%), whereas at Site 2 the structured field was the more complete source (50.4% vs 34.7%). The two sites also differed in how often both formats co-occurred: dual-documentation was present in 1,422 encounters (69.2%) at Site 1 but only 519 (20.9%) at Site 2. Single-format documentation followed the opposite pattern, being more common at Site 2, where 734 encounters (29.5%) carried a structured entry only and 344 (13.8%) an unstructured entry only, compared with 228 (11.1%) and 314 (15.3%) at Site 1. Overall, 890 encounters (35.8%) at Site 2 had no documented periodontal diagnosis in either format, versus 91 (4.4%) at Site 1.

**Table 1:** Completeness of periodontal diagnosis documentation, by data source and site.

| **Metric** | **Sub-metric** | **Site 1 (n = 2,055)** | **Site 2 (n = 2,487)** |
|---|---|---|---|
| **Completeness** | Any Structured (S) diagnosis recorded | 1,650 (80.3%) | 1,253 (50.4%) |
|  | Any Unstructured (U) diagnosis recorded | 1,736 (84.5%) | 863 (34.7%) |
|  | Both S + U present | 1,422 (69.2%) | 519 (20.9%) |
|  | Structured only | 228 (11.1%) | 734 (29.5%) |
|  | Unstructured only | 314 (15.3%) | 344 (13.8%) |
|  | Neither format | 91 ( 4.4%) | 890 (35.8%) |

Among encounters in which a diagnosis was documented in both structured and unstructured formats, agreement between the two sources was high but imperfect (Table 2). Site 1 showed greater concordance (raw agreement 0.88; quadratic-weighted κ = 0.86, 95% CI 0.835 to 0.888) than Site 2 (raw agreement 0.77; κ = 0.80, 95% CI 0.749 to 0.846). Pooling the 1,941 dual-documented encounters across both sites, the two formats assigned the same periodontal stage in 1,654 (85.2%) and diverged in 287 (14.8%). Discordance was more often large than small: 164 encounters (8.4%) differed by two or more stages, compared with 123 (6.3%) that differed by a single adjacent stage (Fig. 2). The disagreements were also directional: the structured field assigned a milder stage than the narrative note in 195 of the 287 discordant encounters (67.9%), versus a more severe stage in 92 (32.1%). Thus, when structured and unstructured diagnoses disagreed, the mismatch more often spanned multiple disease stages than a single boundary, and the coded field more often carried the milder of the two stages.

**Figure 2:**
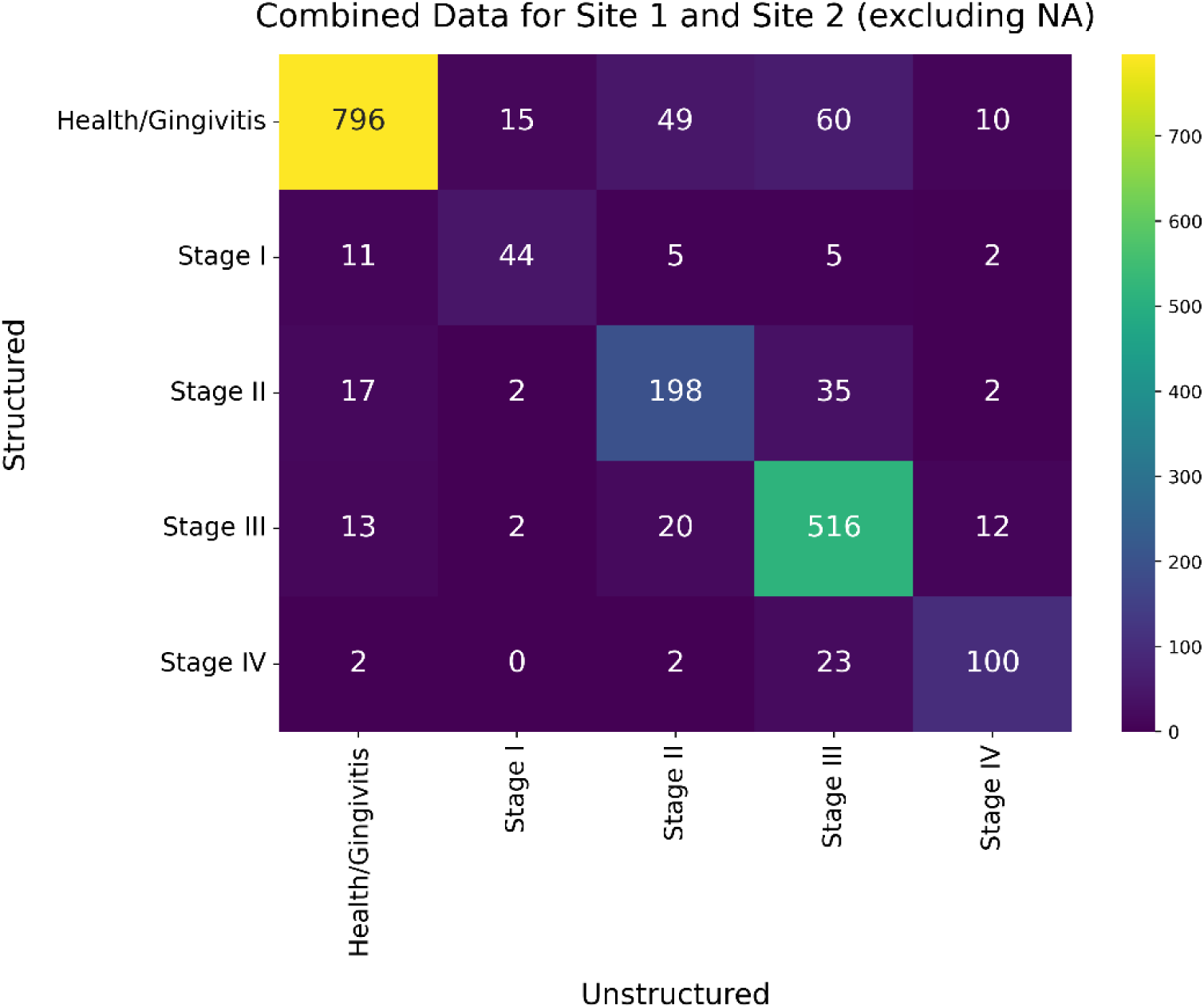
Heatmap of structured vs unstructured diagnoses. Against the expert gold-standard chart review, absolute accuracy was modest for both data sources, and unstructured notes were numerically more accurate than structured fields at both sites (Table 2). At Site 1, structured entries matched the gold standard in 44.5% of cases (n = 472; 95% CI 40.1 to 49.0) versus 50.9% for unstructured diagnoses (n = 407; 95% CI 46.0 to 55.7); the corresponding quadratic-weighted κ values were 0.361 (95% CI 0.280 to 0.442) and 0.509 (95% CI 0.429 to 0.588). At Site 2, structured accuracy was 48.9%(n = 854; 95% CI 45.6 to 52.3) versus 51.6% for unstructured (n = 316; 95% CI 46.1 to 57.0), with weighted κ of 0.320 (95% CI 0.266 to 0.373) and 0.571 (95% CI 0.495 to 0.646). Although unstructured accuracy was higher at both sites, the difference in raw agreement was not statistically significant at either (Site 1: +6.4 percentage points, 95% CI −0.2 to 12.9; Site 2: +2.7 percentage points, 95% CI −3.8 to 9.0). The severity-weighted κ separated the two formats more clearly: the structured and unstructured κ confidence intervals did not overlap at Site 2 and were nearly non-overlapping at Site 1, indicating that the unstructured advantage, though modest in absolute terms, was unlikely to be due to chance at Site 2 and was borderline at Site 1. Overall, absolute accuracy remained modest for both formats, with all point estimates near 50%.

**Table 2:**
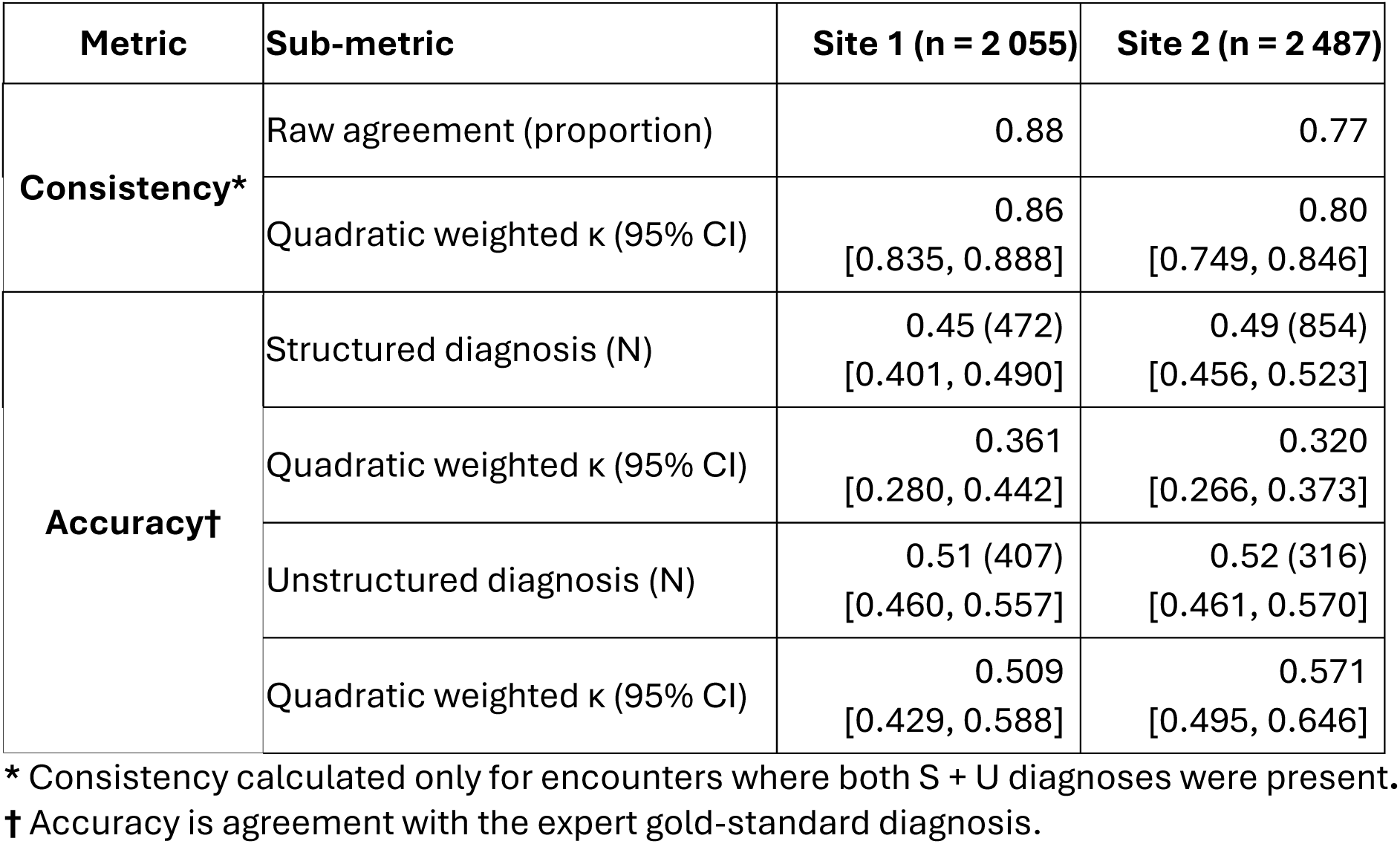
Consistency and accuracy of periodontal diagnoses, by source and site.

## Discussion

Our central question was whether structured (coded) and unstructured (narrative) documentation differ in completeness, accuracy, and consistency, and whether any such differences recur across sites. Three findings stand out. First, neither format was consistently more complete: which one more often carried a diagnosis varied by site, so the documentation discipline imposed by structured templates did not translate into a reliable completeness advantage. Second, two patterns did recur across both independent settings. Both structured and unstructured diagnoses matched the expert gold standard only about half the time, and when both formats were present they agreed substantially but imperfectly. Against this shared backdrop, narrative notes held a small, severity-weighted accuracy edge that was clear at one site and borderline at the other. Together, these results describe a hybrid data asset that is neither reliably complete nor consistently correct. Yet this is the very record a learning health system would draw on to generate knowledge and return it to the point of care. To the best of our knowledge, this is the first comparative analysis of structured versus narrative diagnoses within a dental EHR. Because we analyzed the two sites in parallel rather than against each other, our aim was not to rank institutions but to identify shared, systematic patterns.

A primary dimension we assessed was completeness, defined as the presence of a documented periodontal diagnosis among qualifying patients. Contrary to a simple “structured captures more” narrative, the more complete format differed by site: narrative notes carried a diagnosis slightly more often at Site 1, whereas the structured field was the more complete source at Site 2. Thus, no format was consistently more complete. What did converge across sites, and what we regard as the more concerning completeness finding, was outright missingness: over one-third of qualifying encounters at Site 2 had no documented periodontal diagnosis in either format. Where structured fields did outperform notes, as at Site 2, the mechanisms proposed in prior work, such as mandatory fields and guided templates that prompt clinicians to record essential data points, offer a plausible explanation^23,24^. Such prompts can be beneficial in busy clinical settings by reducing inadvertent omissions, but our data indicate they do not guarantee capture and do not confer a uniform completeness advantage across institutions.

In terms of accuracy, unstructured notes, when present, were numerically more accurate than structured fields at both sites, but this directional advantage warrants caution: the raw differences were modest and non-significant at either site, though clearer on the severity-weighted κ. This is consistent with the view that rigid predefined fields lead clinicians to select the “closest fit” rather than the most precise descriptor, or to “checklist fatigue” in which fields are populated without capturing clinical nuance^25,26^. Correspondingly, the coded field more often carried the milder stage when formats disagreed. Crucially, absolute accuracy was modest for both sources (all estimates near 50%), pointing to a challenge in translating clinical assessment into recorded data regardless of format. This may partly reflect the learning curve of the periodontal classification introduced only a few years earlier^27^, an interpretation supported by a contemporaneous study reporting comparable accuracy^28^.

Lastly, we evaluated consistency, defined as agreement between structured and unstructured diagnoses for the same encounter when both were present. Agreement was substantial but imperfect, and lower at Site 2 than at Site 1. Two features of the disagreements are more informative than the overall agreement rate. First, discordance was more often large than small: among dual-documented encounters, more differed by two or more stages than by a single adjacent stage. Second, disagreement was directional; the structured field assigned a milder stage than the narrative note in roughly two-thirds of discordant encounters. In other words, when the two sources diverged, they tended to diverge by a clinically meaningful margin, and the coded field was the milder of the two in about two-thirds of those cases. We did not adjudicate discordant encounters individually against the gold standard, so we cannot attribute these mismatches to either source; given that absolute accuracy was near 50% for both formats, some discordant pairs likely contain no correct value at all. The directional asymmetry is nonetheless consequential: analytics, population health surveillance, and CDS that rely predominantly on coded data will inherit a coded stage that is systematically the milder of the two available records, and because larger stage discrepancies carry disproportionately greater clinical consequence, this skew is a direct threat regardless of which source is nearer the truth.

Current EHR designs often physically separate structured entry points from narrative note sections, which may contribute to clinicians inadvertently misaligning information between the two. While some might argue that redundant dual entry could serve as a data-quality check, the additional burden on clinicians already facing significant workloads is a serious concern that can contribute to burnout^29^. These challenges underscore the need for more integrated and intelligent EHR documentation workflows, interventions at the practice end of the cycle, aimed at the point where knowledge is meant to re-enter care. Future systems could more seamlessly blend structured and unstructured entry. Dynamic templates^30^ could guide clinicians through comprehensive periodontal assessments, incorporating parameters such as pocket depth, clinical attachment level, and radiographic findings, while offering embedded narrative fields within the same interface. Real-time NLP could parse free-text entries, identify key clinical concepts including diagnoses, and flag discrepancies between the narrative and structured fields for immediate reconciliation. This capability speaks directly to our central discordance finding: because coded fields so often diverged from notes, and diverged by two or more stages in more than half of discordant cases, an NLP-driven check is well positioned to surface precisely the mismatches we observed. For example, if a provider types “Stage III, Grade B periodontitis” in a note but selects a milder or less specific code in a dropdown, an alert could surface the conflict at the point of care, where the clinician, who alone has the patient in front of them, can resolve it. Such reconciliation would improve internal consistency directly; whether it also improves accuracy depends on the clinician resolving the conflict correctly, which our data cannot establish. Semantic enrichment of narrative text with standardized terminologies such as SNODDS or SNOMED CT can further bridge narrative richness and computational tractability, improving retrieval and interoperability.

Our study has several strengths, including the inclusion of two large academic dental institutions using the same EHR platform and diagnostic terminology, which allowed a focused comparison of data types. Analyzing the sites in parallel rather than against each other let us distinguish systematic patterns that recurred across settings (modest accuracy, directional discordance) from site-specific ones (which format was more complete), a distinction that matters for any institution deciding whether these risks are ones it can engineer around locally or ones inherent to the documentation formats themselves. The use of experienced, board-certified periodontists for the gold-standard review and the application of NLP methodology to systematically analyze unstructured text add further rigor.

We acknowledge several limitations. First, this was a retrospective study and thus reliant on the quality and completeness of existing documentation. Second, our findings are from academic dental institutions, and their generalizability to private practice or to institutions using different EHR systems may be limited; relatedly, the direction of the completeness difference diverged by site, so our data do not support a general claim about which format is more complete. Third, the use case was specific to periodontal diagnoses, and data-quality characteristics might differ for other dental or medical conditions. Relatedly, discordant encounters were not individually adjudicated against the gold standard, so we can describe the magnitude and direction of structured–narrative disagreement but cannot say which format was correct in any given mismatch. Fourth, because each format’s accuracy could be scored only where that format carried a diagnosis, the two estimates rested on overlapping rather than identical subsets of the same adjudicated charts and were compared as unpaired proportions. Combined with raw differences that did not reach significance at either site, the narrative advantage should be read as suggestive rather than definitive. Fifth, although our NLP model was fine-tuned and evaluated, all NLP models have inherent limitations in interpreting complex clinical narratives, and the use of synthetic data for fine-tuning, while useful for augmenting training data, may introduce its own biases. Finally, the gold-standard review, though robust, was based on a sample of the data, and expert interpretation retains a degree of subjectivity.

The distinct advantages and limitations of structured and unstructured EHR data necessitate a balanced approach. Structured data offer consistency in format and computability crucial for CDS and large-scale analytics, while unstructured narratives provide contextual richness often missing from coded entries. Our findings suggest that relying solely on one data type may yield an incomplete or inaccurate understanding of patient conditions. Hybrid strategies that leverage the strengths of both, supported by advanced informatics techniques such as NLP and intelligent EHR design, are therefore essential. Combining unstructured with structured data has already been shown to improve prediction-model performance relative to using either source alone^31^.

In two dental centers, both structured and unstructured diagnoses matched expert adjudication only about half the time, and neither format was consistently more complete, with up to one-third of qualifying encounters at one site missing any documented diagnosis. Narrative notes held a small severity-weighted accuracy advantage, and roughly one in seven dual-documented encounters (14.8%) held discordant diagnoses, with 8.4% differing by two or more stages and the coded field carrying the milder stage in 67.9% of disagreements. Improving data quality will require interfaces that marry the computability of coded entries with the fidelity of free text, ideally through integrated templates, real-time NLP, and, eventually, ambient AI documentation support. Until then, a dental learning health system will be learning from a record that is systematically milder than the disease in front of the clinician.

## Data Availability

The data underlying this study are not publicly available. They consist of electronic health record data, including free-text clinical notes, from patients treated at two academic dental institutions, and may contain information that could identify patients.

## Acknowledgements

Funding was provided by the National Institute for Dental and Craniofacial Research (NIDCR) R56DE034086; and the Agency for Healthcare Research and Quality (AHRQ) R01HS027938.

